# Charred or Fragmented, Yet Comparable: Quantifying Dental Surface Similarity Across Teeth, Jaws, and Heat Exposure

**DOI:** 10.1101/2025.04.07.25325362

**Authors:** Anika Kofod Petersen, Scheila Mânica, Andrew Forgie, Richard Boyle, Hemlata Pandey, Palle Villesen, Line Staun Larsen

**Affiliations:** Department of Forensic Medicine, Aarhus University, Denmark; Centre for Forensic and Legal Medicine and Dentistry, University of Dundee, UK; School of Medicine, Dentistry and Nursing, University of Glasgow, Scotland; School of Dentistry, University of Dundee, UK; Bioinformatics Research Centre, Aarhus University, Denmark; Department of Clinical Medicine, Aarhus University, Denmark; Department of Dentistry and Oral Health, Aarhus University, Denmark

**Keywords:** Forensic Odontology Identification, Disaster Victim Identification, 3D Dental Comparison, Biometric Identification

## Abstract

Accurate dental matching is essential for forensic identification, particularly in challenging cases involving dentitions with no dental work, incomplete dentitions or damaged remains. This study evaluates similarity scoring schemes for 3D dental data using three datasets: full jaws versus single teeth (DATA-A), and two collections of heat-traumatized teeth (DATA-B and DATA-C). The similarity scores are assessed for their ability to quantify curvature similarity and distinguish matching from mismatching dental comparisons. Results demonstrate the method’s effectiveness in handling dental fragmentation (ROC-AUC_DATA-A_ = 0.899 (95% CI 0.840 to 0.948) and heat trauma (ROC-AUC _DATA-B_ = 0.996 (95% CI 0.98 to 1.00); ROC-AUC _DATA-C_ = 0.993 (95% CI 0.980 to 1.00), offering a robust tool for forensic applications.

**Highlights:**

1. Tests the keypoint pipeline on two different trauma scenarios
2. Quantifies dental surface similarity between single teeth and full jaws
3. Quantifies dental surface similarity before and after heat exposure
4. Underlining the usefulness of the keypoint pipeline for trauma scenarios

## 1. Introduction

Disaster Victim Identification (DVI) is essential for providing legal certainty and individual rights in the case of mass casualty events [1-5]. Mass casualty events, such as natural disasters, resulting in numerous fatalities, require substantial resources to manage their aftermath [4,5]. To minimize the impact and support to both families and authorities, DVI must be swift, efficient, and ensure accurate identification [2-7]. Classic forensic odontology identification is time consuming, as full *post mortem* dental examinations must be compared to *ante mortem* dental records [1-4,6-8]. To aid in the identification process, one broadly used software is the KMD PlassData DVI (Disaster Victim Identification), currently acquired by 31 countries [6,9]. This software, and the INTERPOL guideline [3], encourages the use of dental work, i.e. fillings, crowns, implants and root canal treatments, for decision of identity [2,3,7,9-11]. When no dental work is present in the *post mortem* dentition, the way of identification is a thorough *post mortem* examination focusing on the distinctive dental anatomy and abnormalities of the teeth and malocclusions, followed by comparison with *ante mortem* dental records [2,3,5,7,8,10,11]. This task is cumbersome, and with a rise in dental health in many countries worldwide, forensic odontology identification would benefit from a quantitative similarity measure which is not dependent on dental work and that can be semi- or fully automated [4,7,12,13].

Our research group has previously proposed a pipeline for quantifying dental surface similarity based on intraoral 3D scans and a similarity score to distinguish between pairs of scans (e.g. a query scan of a *post mortem dentition* and a target scan from a database of *ante mortem* dentitions) [14-16]. Our approach has been tested on dental data with 6 months’ time difference, and full and partial dentitions with as little as a quarter of the scanned oral surface overlapping [15,16]. The similarity score is based on matching curvature keypoints on the dentition surface between two scans and ranges from 0 (low similarity) to 1 (identical) [16].

Even though the approach has been tested on data with a time difference and partial dental data, it has yet to be tested on more extreme cases, mimicking disaster conditions, such as single teeth and burned teeth [15,16]. *Ante mortem* intraoral 3D scans would, if possible, cover the entire dentition surface of both the upper and the lower jaw. But for *post mortem* 3D scans, as little as a single tooth might be recovered, and the PM dentition might be damaged by external factors such as burning, which can harm the dental surface [5,8]. In cases with little dental material and/or trauma to the dental hard tissue affecting the surface, becomes especially challenging [2,3,5,6,8].

This study aims to test the similarity score approach in challenging simulated disaster settings and compare it the previously suggested scoring scheme. First the ability to identify matches of single *post mortem* teeth to full *ante mortem* jaws is tested, followed by comparison of single teeth before and after heat trauma.

## 2. Materials and Methods

The approach consists of 4 steps [15,16]. First, extreme curvatures of the dentition (all dentition surfaces) are located [15]. Secondly, the dentition surface curvatures within a 2 mm radius of a keypoint are represented in a multidimensional space [15]. Thirdly, all keypoint representations from a query scan are compared with all keypoint representations from a target scan (typically from a database with many *antemortem* scans) [15,16]. Lastly, similarity is calculated from the arithmetic mean of the 15 most similar keypoints and the General Procrustes Analysis (GPA) disparity of the 8 best matching keypoints as a weighted average (90%/10% respectively) [16]. In this study, we also compare to a previous scoring scheme [15].

### 2.1 Data

We worked on three different datasets. Dataset A (DATA-A) consists of AM full jaws and PM single teeth. Dataset B and C (DATA-B, DATA-C), collected from two different study centres, consist of single teeth before and after exposure to heat trauma. All IOS in this study were stored as meshes in the STL file format.

### 2.2 Dataset A

DATA-A consisted of a query set (89 single teeth), and a target set (102 full jaws). The 102 full jaw scans (upper and lower jaws from 51 individuals) were all pre-processed using grid-cutting [14]. The 89 single teeth were manually cut from full jaw scans from 6 of the 51 individuals who were scanned again after approximately 6 months.

After keypoint detection of the single teeth, 3 teeth were discarded (one molar (M3), one canine and one premolar) since less than 15 keypoints were identified on the dental surface. Of the 8772 query-target comparisons (86 teeth*102 jaws) there were 86 matches and 8,686 mismatches.

Since the single teeth were cut from full jaws, they did not contain entire proximal dental surfaces and associated soft tissue. This means that the scan border in some cases was too close to the occlusal surface, which was expected to harbour much of the dental surface curvature (Figure 1). It should be noted that due to the calculation instability on surface borders, keypoints were not allowed to be found within a 2 mm distance from the border of a scan, thus lowering the number of keypoints found on the single teeth dental surfaces. This was expected to be the major cause of exclusion for this dataset, alongside a flat curvature landscape.

**Figure 1.**
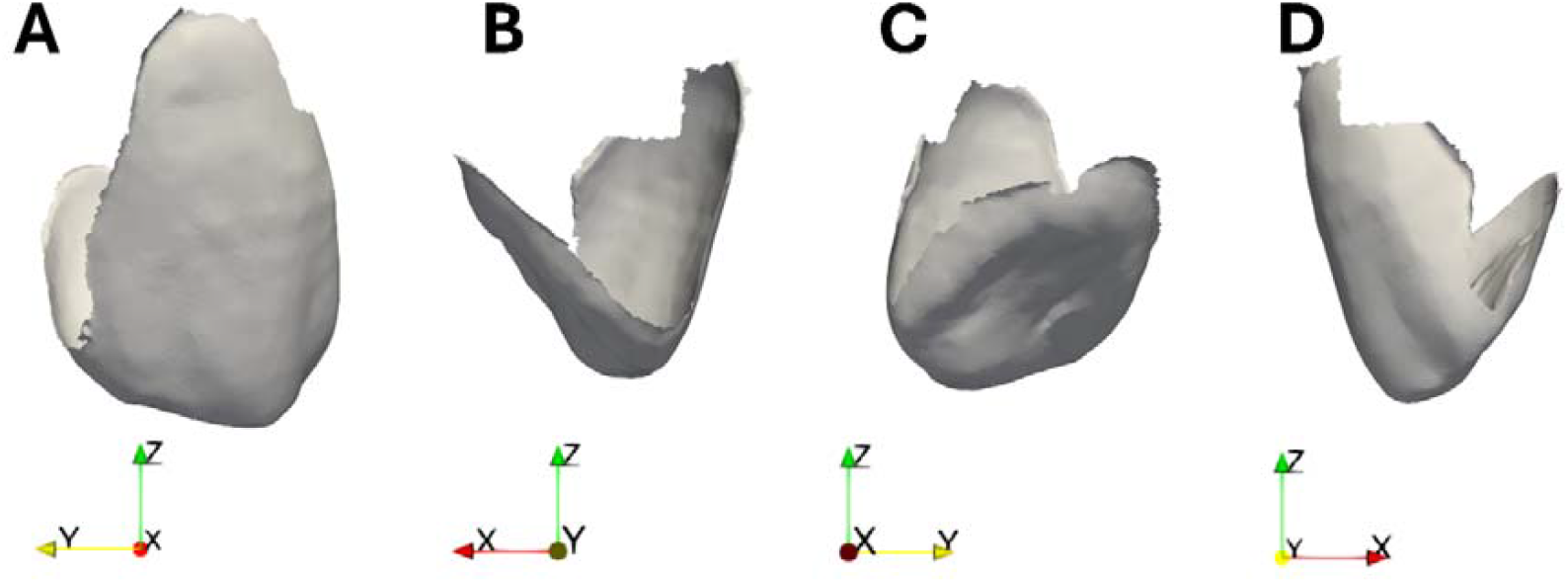
A canine tooth cut from a full jaw scan showcasing the lack of proximal dental surface. The tooth is shown in A) facial view B) distal view C) lingual view and D) mesial view

### 2.3 Dataset B and Dataset C

DATA-B and DATA-C constituted two similar datasets: one originating from Aarhus University and the other originating from the University of Dundee. Both datasets comprised scans of 30 extracted permanent teeth mounted individually in plaster of Paris and scanned before and after exposure to heat trauma. Detailed specifications for the two datasets from the different study centres are listed in Table 1.

**Table 1:** Methodological details considering the laboratory part of DATA-B and DATA-C.

|  | Aarhus University | Remarks | University of Dundee | Remarks |
| --- | --- | --- | --- | --- |
| Specimen origin | The specimens were drawn from an anonymous tooth biobank (the Central Denmark Region Committees on Health Research Ethics, case No. 1-10-72-1-21) holding donated teeth for educational and research purposes at the Department of Dentistry and Oral Health, Aarhus University. | Until inclusion, the teeth were stored in water with thymol (0.4 %). The teeth were kept moist during the experiment until heat treatment. | The specimens belonged to the EDIMOS tooth biobank pre-2006. The Human Tissue (Scotland) Act 2006 regulates the use, storage, and disposal of human tissue, organs, and bodies. Collections existing prior to 1st September 2006 can be used for educational and research purposes without patient consent respecting local policies. | The teeth were stored in plastic containers with phosphate-buffered saline (PBS) solution (pH ≈ 7.4). The teeth were kept moist during the experiment until heat treatment. |
| Plaster of Paris | NEO DURODERM® 7, Hera, KULZER, Germany |  | GC FUJIVEST SUPER®, GC Corporation, Tokyo, Japan |  |
| Scanner | Primescan AC, Sirona Dental Systems GmbH, Germany |  | Primescan AC, Sirona Dental Systems GmbH, Germany |  |
| Furnace | Magma, 220-240 V, Renfert, GmbH, Germany | The oven was pre-heated to a definite temperature before inserting the specimens. | Carbolite CSF 1200, Carbolite Gero Ltd., Hope Valley, UK | The oven was pre-heated to a definite temperature before inserting the specimens. |

Figure 2 shows a chart of the teeth included and the experimental conditions for each dataset. Extracted teeth were drawn from two local tooth biobanks at the respective study centres and mounted in plaster of Paris. The target scans were compiled by 3D scanning each tooth crown prior to any trauma. Then the teeth were burned at 300 degrees Celsius in a furnace for either 15 or 30 minutes, left at room temperature to cool down and then 3D scanned again (the query scans). Exclusion criteria of teeth comprised: gross pathology with distinct tissue loss, e.g. severe abrasion/erosion, obvious fractures, and prosthetic tooth crowns. Both unrestored (no abnormality detected) and teeth restored with composite or silver amalgam were included (see Figure 2).

**Figure 2.**
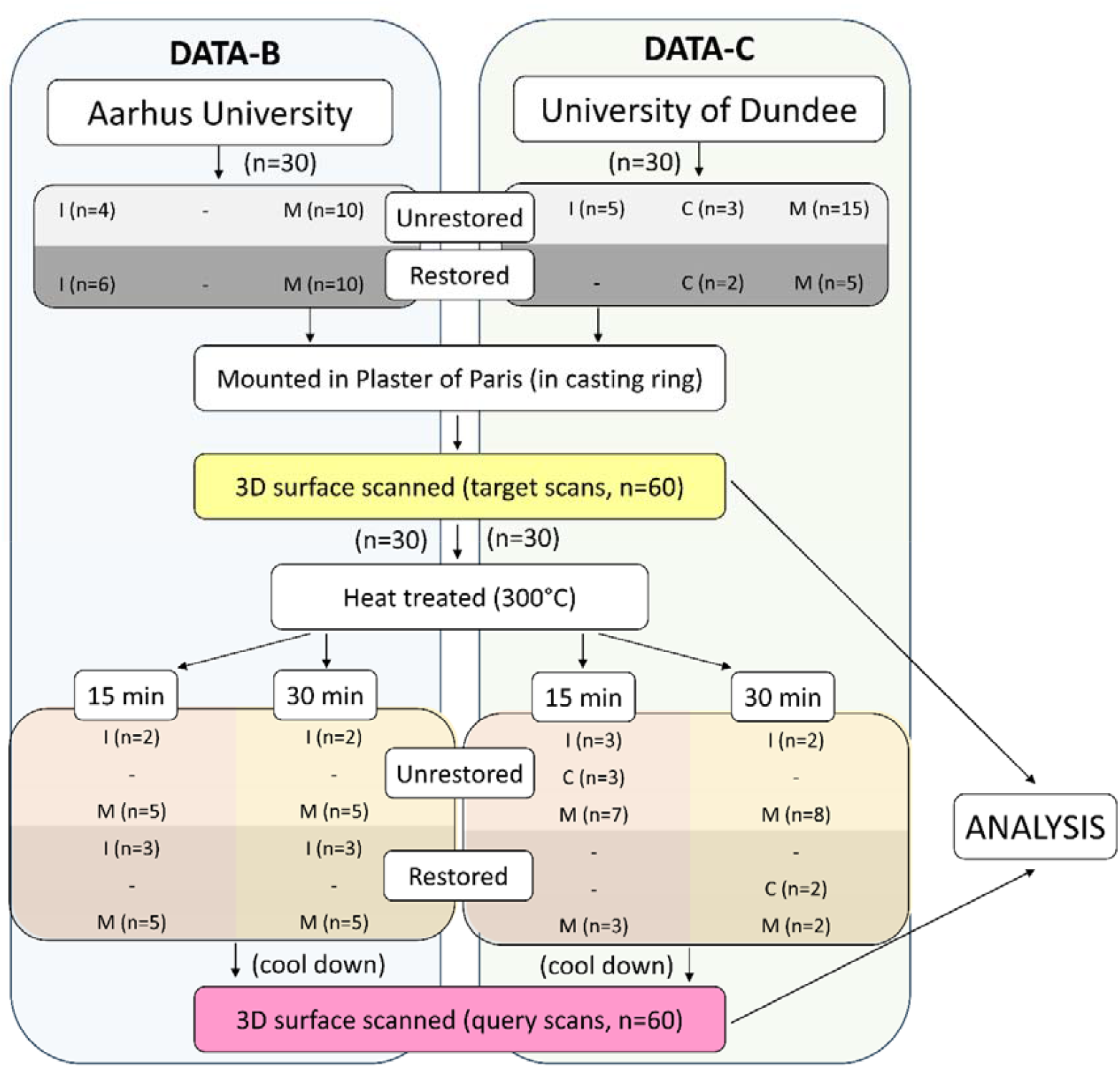
Schematic diagram of the laboratory part of DATA-B and DATA-C.

Experimental time intervals and degree measures were decided after running and discussing three pilot studies conducted at University of Dundee and Aarhus University, respectively. Due to the conditions of the scientific method we aimed to test, based on tooth surfaces, it was important that the teeth did not break or splinter completely during the heat treatment. Before analysis, all teeth from DATA-B and DATA-C were cropped to exclude the plaster of Paris from the mesh surface. Each of the two datasets hold 30 matches and 870 mismatches.

All three datasets were kept separate and treated as if they were from different disasters, i.e. analysed independently. As explained previously, all datasets were pre-processed by limiting non-dental surfaces, either by grid-cutting or by manual removal of the plaster of Paris from the scans. All scans were subjected to keypoint detection and keypoint representation as outlined in our previous work. For comparison, all the keypoints from each query scan were compared to all the keypoints from each target scan, thus giving every comparison a similarity score. Each comparison was labelled either as a match, if the dental surfaces in the two scans originated from the same dentition (same jaw and same individual), or as a mismatch if the two scans did not originate from the same dentition.

To account for the class imbalance in the dataset, all datasets were subjected to stratified bootstrapping with random under-sampling of the majority class. This was done for 1,000 bootstraps. The receiver operating characteristic (ROC) area under the curve (AUC) was investigated by comparing the score given to a query-target comparison with the true label (either match or mismatch) for every bootstrap, with the AUC being the mean AUC across bootstraps [17-19]. The 95% confidence interval (CI) was calculated based on the ROC-AUC for each bootstrap.

## 3. Results

### 3.1 Single teeth matching against full jaws (DATA-A)

When comparing the single teeth with the full jaws from DATA-A, we found an excellent performance (AUC = 0.899, 95% CI 0.840 - 0.948) (Figure 3). The combined score and the arithmetic mean alone gave almost the same performance on DATA-A, while outperforming the IQR score and GPA disparity. The score distributions of the matching and mismatching classes can be seen in supplementary materials Figure 1. A Kruskal-Wallis test showed no significant difference between tooth types (p = 0.211), and Mann-Whitney U tests between pairs of tooth types also showed no significant difference (p>0.05) (Figure 4A). When investigating how well matches rank compared to mismatches, in the majority of cases the true match was the best ranking result (Figure 4B). Some tooth types had a tendency towards better ranking, such as canines that showed 90.9% of cases where the true match was the best scoring comparison. There was a significant difference between ranks of the different tooth types (Kruskal-Wallis, p=0.026). Between pairs of tooth type ranks, there was a significant difference (p<0.05) when comparing incisors and molars (M1, M2), incisors and canines, and canines and wisdom teeth (M3) (Figure 4B).

**Figure 3.**
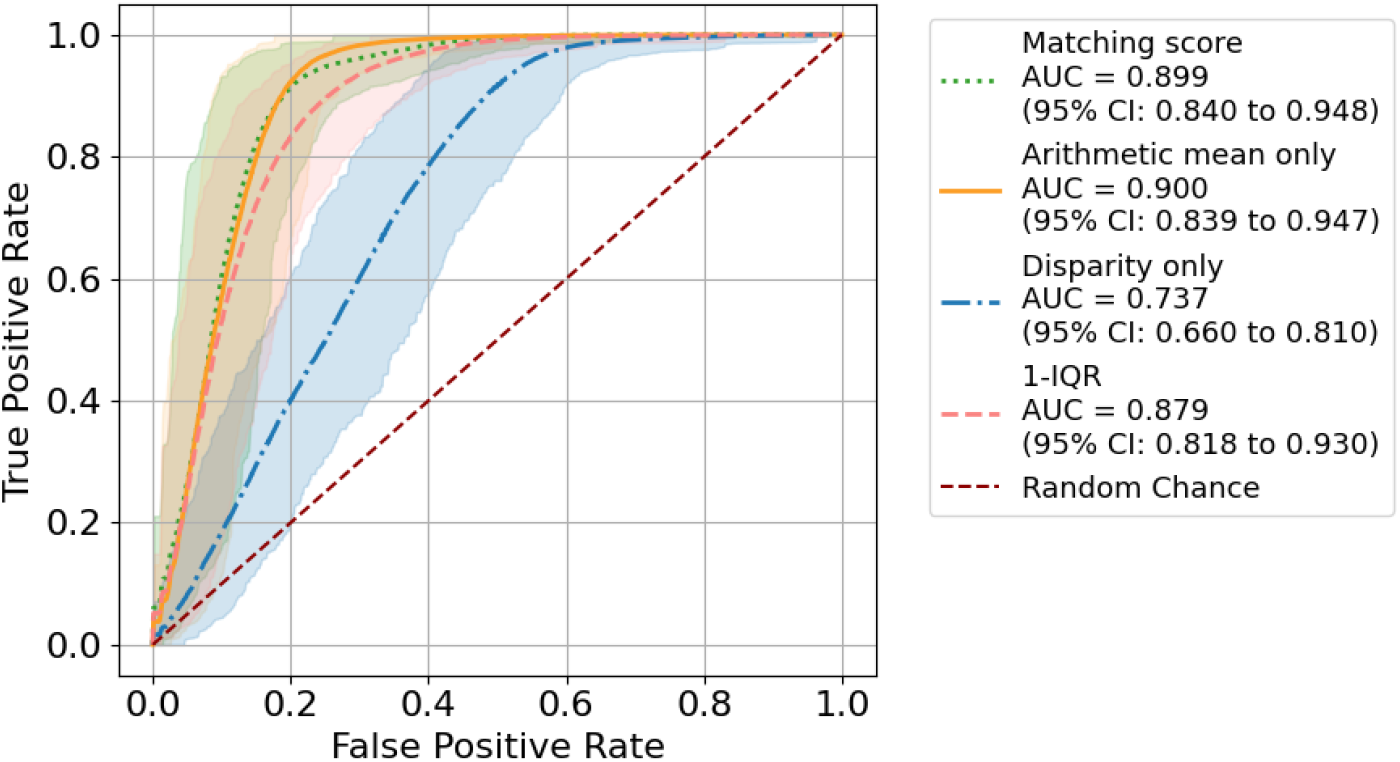
ROC-AUC of the comparisons of AM full jaws vs single teeth from DATA-A based on the four scoring schemes

**Figure 4.**
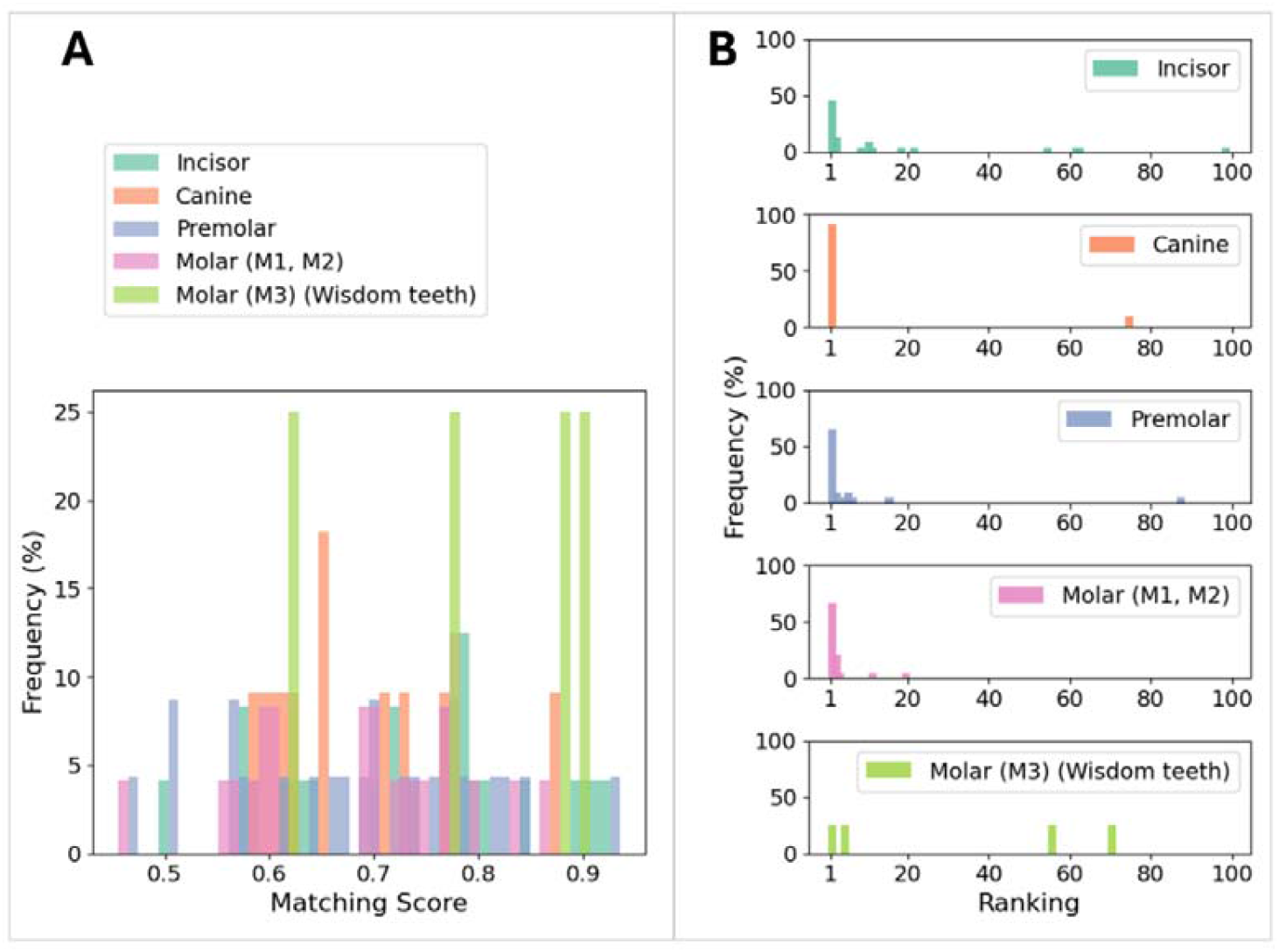
Scoring of comparisons using the favoured scoring scheme showing A) the raw scores of the 86 matches and B) the ranked scores based on tooth type.

### 3.2 Heat traumatized teeth

When comparing single teeth before and after heat traumatization we found an even higher discriminatory performance for both DATA-B (teeth from Aarhus university, Figure 5) and DATA-C (teeth from university of Dundee, Figure 6). The high discriminatory performance is comparable with the previously reported performance for comparison of full and partial jaws [15], but with a larger confidence interval. The score distributions of the matching and mismatching classes can be seen in supplementary Figure 2. For DATA-B, no significant difference was seen between raw scores given to molars and incisors (Mann-Whitney U test, p=0.375) (Figure 5A). For DATA-C, a significant difference was seen between the three tooth types (Kruskal-Wallis test, p<0.001) (Figure 6A). For the individual tooth types in DATA-C, only incisor and canines didn’t show significant difference in scores (Mann-Whitney U test, p=0.970).

**Figure 5.**
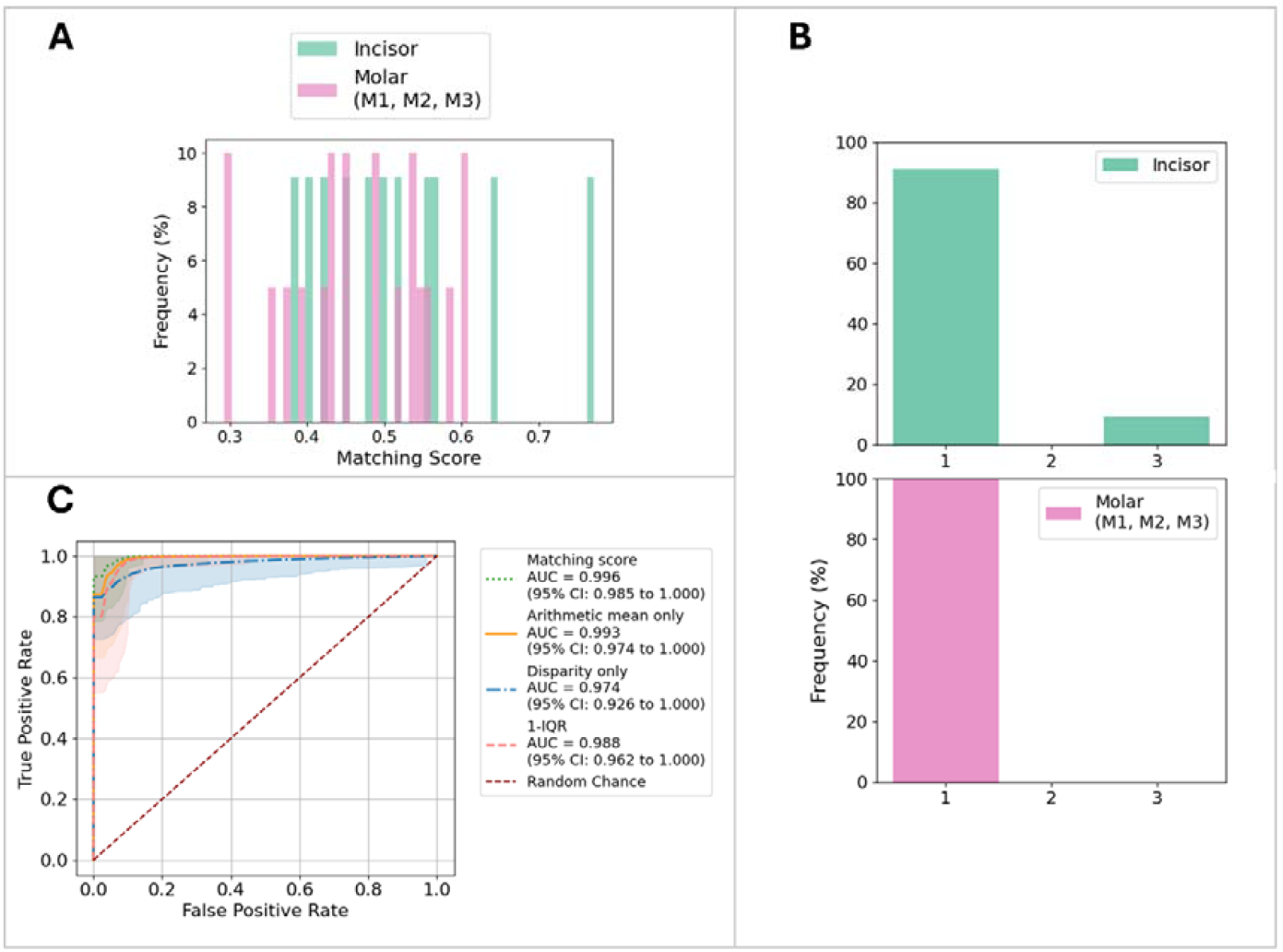
Scoring of DATA-B comparisons using the favoured scoring scheme showing A) the raw scores, B) the ranked scores based on tooth type, C) ROC-AUC based on the four scoring schemes

**Figure 6.**
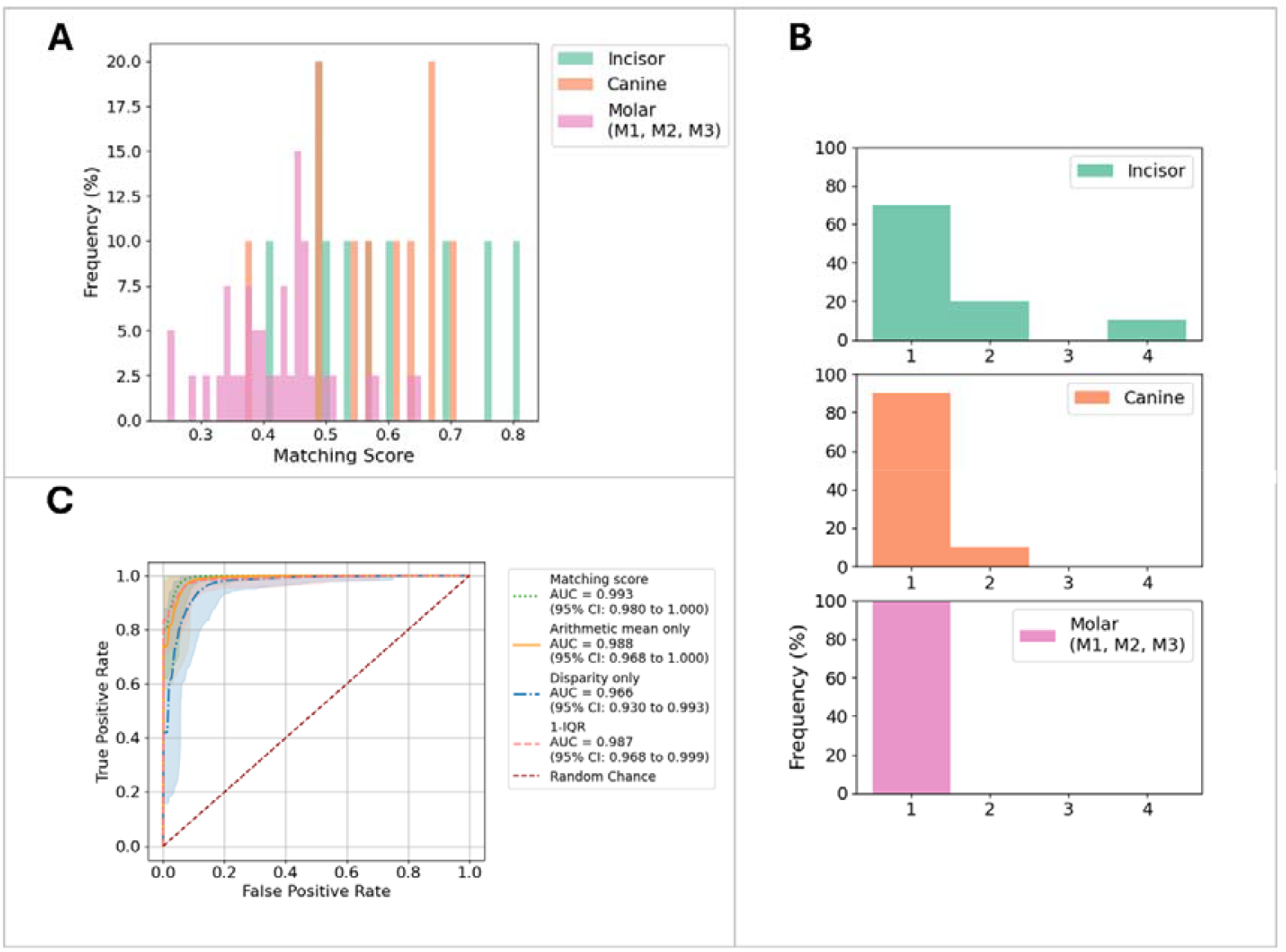
Scoring of DATA-C comparisons using the favoured scoring scheme showing A) the raw scores, B) the ranked scores based on tooth type, C) ROC-AUC based on the four scoring schemes

Looking at the ranking for DATA-B, we see no significant difference between tooth type groups (Mann-Whitney U test, p=0.200). For DATA-C, there was a significant difference between tooth type groups (Kruskal-Wallis, p=0.003). Incisors and molars showed pairwise significant difference in ranking using Mann-Whitney U test (p<0.001).

The high discriminatory performance is also evident in the ranking analysis. For DATA-B only one true match was not the best scoring comparison and was out ranked by two other comparisons (Figure 5B). Upon investigation it was found that for this comparison the tooth fractured while in the furnace, thus potentially altering keypoint detection and keypoint representation. For DATA-C, four true match comparisons were out ranked by mismatch comparisons (Figure 6B). Two of these true match comparisons involved a tooth that fractured during heat traumatization, resulting in ranking fourth and second.

## 4. Discussion and conclusions

In this study, we tested the performance of previously proposed scoring schemes for dental surface similarity [15,16]. We see that even in the cases of single teeth and heat traumatized teeth the scoring scheme can give a quantitative indication of matching status.

This study shows that the comparison of single heat traumatized teeth has better discrimination between matches and mismatches than when comparing single teeth to full jaws [15,16]. This result can potentially emerge from two main causes. First, in the data acquisition and preprocessing phase for the single teeth in DATA-A, much of each tooth’s dental surfaces are removed, limiting the available dental surface for keypoint detection. Especially when considering the limitation of distance to the intraoral 3D scan border. Secondly, keypoints are detected on surfaces of different scale (single teeth vs full jaws) which might impact the keypoints chosen, since keypoints are found based on relative curvature extremes. Therefore, it might be possible to increase performance, if the full jaw was subject to segmentation, and each tooth where matched one against another. This second possible causation is deemed unlikely, as previous studies have shown that matching a quarter of the scanned oral surface to the full scanned oral surface did not suffer from this [15].

The burned teeth of DATA-B and DATA-C were compared on similar surface scales, both being single tooth scales. Here we see similar performance as for matching on larger surface areas, but with an increase in confidence intervals [16]. This is much in line with the logic that a larger matching surface would increase the confidence in calling a match for a forensic odontologist. Furthermore, we found that for DATA-C molars were easier to correctly match than incisors. It was expected that this would be true for DATA-A and DATA-B as well, due to a more distinct curvature landscape of molars. But no significant difference was shown for these two datasets. Third molars (M3, wisdom teeth) from DATA-A showed to be difficult to recognise (Figure 4B). This could be caused by M3s from DATA-A being cut from IOS, causing loss of the proximal surfaces, loss of surrounding soft tissue and adjacent teeth, resulting in a different curvature landscape. Since the third molars are positioned at the back of the mouth, and in some cases have only partly erupted, they tend to be more difficult to scan than other teeth. This could also add to the inaccuracy in query-target matching.

By testing the scoring schemes on these challenging simulated disaster datasets, a broader understanding of how they can automate ranking of *ante mortem* dental records, ultimately increase efficiency in forensic odontology identification has been established, especially in cases where traditional identification would be challenging [2,5,6,8,10].

In practice, the matching scoring scheme and the matching pipeline could be incorporated into disaster victim identification software [6,9], allowing the forensic odontologist to gain insights on matching likelihood of the 3D dental surface, especially in cases where filtering the *ante mortem* database based on dental work is of little use [2,3,7,8,11,13]. This study underlines that even in the case where single teeth are recovered *post mortem*, the matching pipeline can still be of use [15,16].

## Data Availability

The research data consists of personal dental data that the authors are not authorized to share

## Acknowledgements

We thank AUFF NOVA for funding this study (Grant number: AUFF-E-2021 -9-14). Part of the computing for this study was carried out on the GenomeDK cluster. We thank GenomeDK and Aarhus University for providing computational resources and support for this research.

## Supplementary material

**Figure 1.**
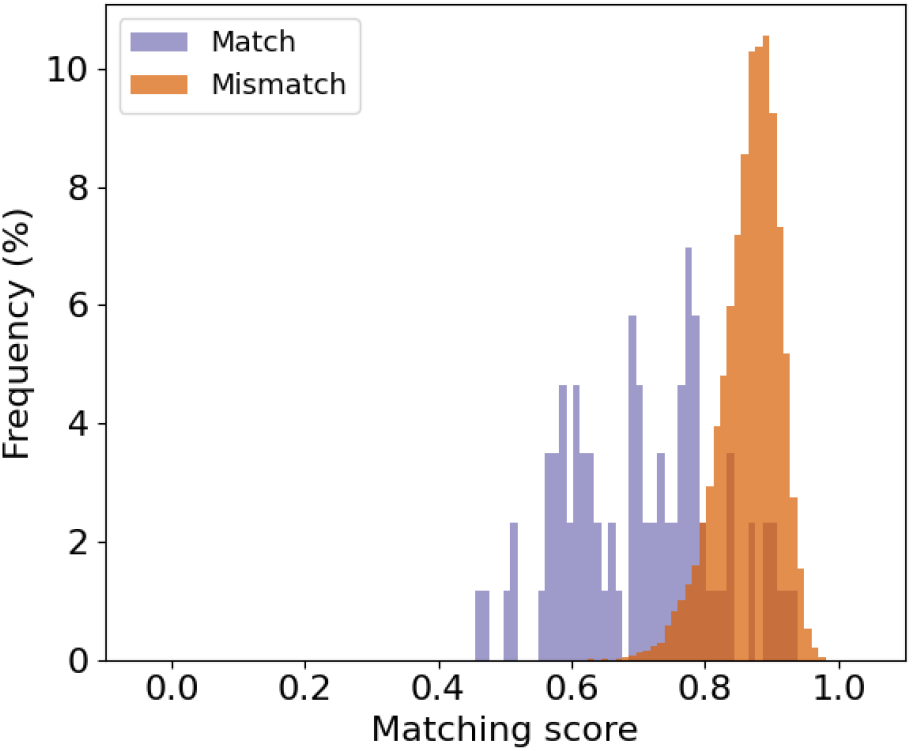
Raw scores of the favoured scoring scheme of matches and mismatches from DATA-A

**Figure 2.**
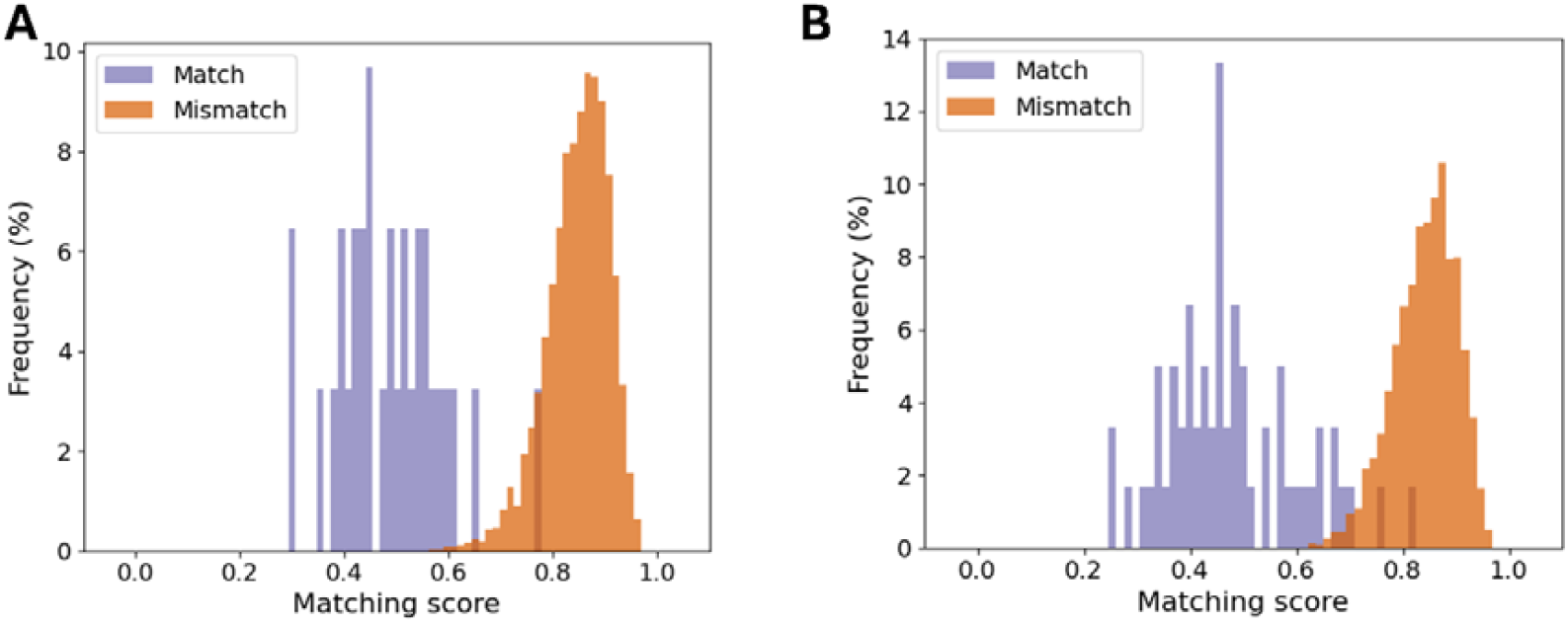
Raw scores of the favoured scoring scheme of matches and mismatches from A) DATA-B and B) DATA-C

## References

1 Berketa, J. W., James, H. & Lake, A. W. Forensic odontology involvement in disaster victim identification. Forensic Science, Medicine, and Pathology 8, 148–156 (2012). 10.1007/s12024-011-9279-9

2 Interpol. Disaster Victim Identification (DVI) https://www.interpol.int/en/How-we-work/Forensics/Disaster-Victim-Identification-DVI, (accessed 19th February 2025). (2025).

3 Interpol. in Part B, Annexure 8: Methods of Identification (November 2023).

4 Jain, P. Mass Fatality Incidence and Disaster Victim Identification-A Comprehensive Review. IRJET 11, 440–450 (2024).

5 de Boer, H. H., Roberts, J., Delabarde, T., Mundorff, A. Z. & Blau, S. Disaster Victim Identification Operations with Fragmented, Burnt, or Commingled Remains: Experience-Based Recommendations. Forensic Sciences Research 5, 191–201 (2020). 10.1080/20961790.2020.1751385

6 Sweet O.C, D. INTERPOL DVI best-practice standards—An overview. Forensic Science International 201, 18–21 (2010). 10.1016/j.forsciint.2010.02.031

7 Sweet, D. Forensic dental identification. Forensic Science International 201, 3–4 (2010). 10.1016/j.forsciint.2010.02.030

8 Kvaal, S. I. Collection of post mortem data: DVI protocols and quality assurance. Forensic Science International 159, S12–S14 (2006). 10.1016/j.forsciint.2006.02.003

9 KMD. Computerized Identification of Disaster Victims & Missing Persons (https://www.kmd.net/solutions-and-services/data-and-ai/kmd-plassdata-dvi), Accessed 5th Feb 2025).

10 Brough, A. L., Morgan, B. & Rutty, G. N. The basics of disaster victim identification. Journal of Forensic Radiology and Imaging 3, 29–37 (2015). 10.1016/j.jofri.2015.01.002

11 Forrest, A. Forensic odontology in DVI: current practice and recent advances. Forensic Sciences Research 4, 316–330 (2019). 10.1080/20961790.2019.1678710, note = PMID: 32002490

12 Biedermann, A., Bozza, S., Taroni, F. & Garbolino, P. A formal approach to qualifying and quantifying the ‘goodness’ of forensic identification decisions. Law, Probability and Risk 17, 295–310 (2018). 10.1093/lpr/mgy016

13 Perkins, H., Hughes, T., Forrest, A. & Higgins, D. 3D dental records in Australian dental practice – a hidden gold mine for forensic identification. Australian Journal of Forensic Sciences 0, 1–-18 (2024). 10.1080/00450618.2024.2359432

14 Kofod Petersen, A., Forgie, A., Bindslev, D. A., Villesen, P. & Staun Larsen, L. Automatic removal of soft tissue from 3D dental photo scans; an important step in automating future forensic odontology identification. Scientific Reports 14, 12421 (2024). 10.1038/s41598-024-63198-2

15 Kofod Petersen, A., Forgie, A., Villesen, P. & Larsen, L. S. 3D Dental Similarity Quantification in Forensic Odontology Identification. Forensic Science International, 112462 (2025). 10.1016/j.forsciint.2025.112462

16 Kofod Petersen, A., Arenholt Bindslev, D., Forgie, A., Villesen, P. & Staun Larsen, L. Objective comparison of 3D dental scans in forensic odontology identification. medRxiv, 2025.2003.2031.25324929 (2025). 10.1101/2025.03.31.25324929

17 Hasanin, T. & Khoshgoftaar, T. The Effects of Random Undersampling with Simulated Class Imbalance for Big Data. 2018 IEEE International Conference on Information Reuse and Integration (IRI), 70–79 (2018). 10.1109/IRI.2018.00018

18 Shankar, P. M. Tutorial overview of simple, stratified, and parametric bootstrapping. Engineering Reports 2, e12096 (2020). 10.1002/eng2.12096

19 Fawcett, T. An introduction to ROC analysis. Pattern Recognition Letters 27, 861–874 (2006). 10.1016/j.patrec.2005.10.010

